# Diverse Humoral Immune Responses in Younger and Older Adult COVID-19 Patients

**DOI:** 10.1101/2021.01.12.21249702

**Authors:** Jennifer M. Sasson, Joseph J. Campo, Rebecca M. Carpenter, Mary K. Young, Arlo Z. Randall, Krista Trappl-Kimmons, Amit Oberai, Christopher Hung, Joshua Edgar, Andy A. Teng, Jozelyn V. Pablo, Xiaowu Liang, Angela Yee, William A. Petri, David Camerini

## Abstract

We sought to discover links between antibody responses to SARS-CoV-2 and patient clinical variables, cytokine profiles and antibodies to endemic coronaviruses. Serum from patients of varying ages and clinical severity were collected and used to probe a novel multi-coronavirus protein microarray containing SARS-CoV-2 proteins and overlapping protein fragments of varying length as well as SARS-CoV, MERS-CoV, HCoV-OC43 and HCoV-NL63 proteins. IgG, IgA and IgM antibody responses to specific epitopes within the spike (S), nucleocapsid (N) and membrane proteins (M) were higher in older adult patients. Moreover, the older age group displayed more consistent correlations of antibody reactivity with systemic cytokine and chemokine responses when compared to the younger adult group. A subset of patients, however, had little or no response to SARS-CoV-2 antigens and disproportionately severe clinical outcomes. Further characterization of these serosilent individuals with cytokine analysis revealed significant differences in IL-10, IL-15, IP-10, EGF and sCD40L levels when compared to seroreactive patients in the cohort.

## Introduction

With cases continuing to rise in the United States as we pass the annual mark since the beginning of the pandemic, the scientific community continues to strive to further characterize the immune response to SAR-CoV-2 infection. COVID-19 leads to a wide range of clinical responses, varying from minor symptoms, effective immune response and viral clearance to major respiratory compromise, significantly uncoordinated immune response and subsequent death (1). Defining antibody responses, both qualitatively and quantitatively, is necessary in characterizing illness severity, assessing treatment strategies and understanding long-term protection after vaccine administration.

The antibody response to SARS-CoV-2 infection consists of a rise in immunoglobulin-M (IgM), and a simultaneous or nearly synchronous rise in immunoglobulin-G (IgG) within the first 14-20 days of infection, plateauing on average about 6 days after seroconversion (2,3). Antibodies targeting the nucleocapsid (N) protein, a 488 amino acid (aa) SARS-CoV-2 internal structure that functions in compaction and protection of the viral RNA genome, and spike (S) protein, a 1273 aa protein that functions in fusion of viral to host cell membranes by binding to cellular receptors, have been implicated as the dominant antibodies through the course of infection (4–6).

Correlation of antibody levels to severity of disease in previous studies have yielded mixed results, owing to the heterogeneity of immune responses seen in COVID-19 infection (4,7). There is limited data, however, on antibodies to specific epitopes within these viral proteins and their association with disease severity.

A large cohort study consisting of over 17 million patients identified common patient characteristics and comorbidities as predictors of death from COVID-19. Among these, age was found to be the strongest predictor of poor outcome (8). This, therefore, raises the question of the differences in antibody response to infection between age groups. Prior studies have shown that older age is associated with increased antibody response (9). Other studies suggest that older age promotes uncoordinated interactions between the branches of the adaptive immune response which ultimately leads to poor outcomes (10). This suggests that the wide range of clinical presentations of COVID-19 could be attributed to multiple interactions between the components of the adaptive response which are influenced by patient demographics and comorbidities.

Given the consistent circulation of endemic coronaviruses in the population, also known as “common cold” coronaviruses, there is interest in the cross-reactivity of antibodies directed to these viruses with SARS-CoV-2 and their subsequent effect on clinical outcomes of COVID-19 (11). The endemic human coronaviruses (HCoV) include alpha (HCoV-229E and HCoV-NL63) and beta (HCoV-OC43 and HCoV-HKU1) subgroups, with the latter also made up of B (containing SARS-CoV and SARS-CoV-2) and C (containing MERS-CoV) lineages (11).

Whether it be through cross-protection or antibody-dependent enhancement of infection, more studies are needed to determine the immune interaction between responses to endemic coronaviruses and how they affect disease severity from COVID-19.

We sought to fill some of these knowledge gaps through use of a novel multi-coronavirus protein microarray with its ability to identify antibody responses to small epitopes using various sized viral protein fragments of SARS-CoV-2. Serum from COVID-19 patients were exposed to these arrays with subsequent correlation of relevant clinical data collected from medical records. This microarray also allowed for correlation of the antibody response to SARS-CoV-2 to coronaviruses of other subtypes and lineages. Overall, we looked to further characterize specific antibody responses to SARS-CoV-2, their correlation with other known coronaviruses and their association with patient clinical data.

## Results

The multi-coronavirus protein microarray used in this study included four structural proteins and five accessory proteins of SARS-CoV-2 created through coupled *in vitro* transcription and translation (IVTT): S, envelope (E), membrane (M), N, open reading frames (ORF’s) 3a, 6, 7a, 8 and 10. Fragments of these nine proteins were made through IVTT in 50% overlapping segments of 30 aa, 50 aa and 100 aa lengths. There were additional structural proteins from SARS-CoV, MERS-CoV, HCoV-NL63 and HCoV-OC43, plus overlapping 13-20 aa peptides of the SARS-CoV structural proteins and of the S proteins of MERS-CoV, HCoV-NL63 and HCoV-OC43 (Table S1). Serum samples were collected from COVID-19 patients of varying ages and disease severity from April 2020 until July 2020. Thirty of these serum samples were used to probe the multi-coronavirus protein microarray. Clinical data including patient medical history, clinical laboratory assays and clinical course were collected from electronic medical records (Tables 1, S2). Serum samples were additionally analyzed by MILLIPLEX ® SARS-CoV-2 Antigen Panel 1 IgG, IgA and IgM for comparison.

**Table 1.** Clinical characteristics of older and younger COVID-19 patients with older age group stratified by ventilation status.

|  | **Young Adult Patients**<br>**(N=10; Age 27-39)** | **Older Patients, Not Ventilated**<br>**(N=11; Age 69-82)** | **Older Patients, Ventilated**<br>**(N=9; Age 69-83)** |
| --- | --- | --- | --- |
| **Age in years (std. dev.)** | 34.3 (3.86) | 75.2 (4.79) | 72.7 (4.5) |
| **Sex** | 30% Male<br>70% Female | 63.6% Male<br>36.4% Female | 77.8% Male<br>22.2% Female |
| **Days from Symptom Onset (std. dev.)** | 8.80 (4.92) | 14.4 (5.80) | 16.8 (11.4) |
| **Race** | 60% Other<br>20% Caucasian<br>20% Unknown | 27.3% Caucasian<br>63.6% African American<br>9.09% Other | 66.7% Caucasian<br>11.1% African American<br>22.2% Other |
| **Ethnicity** | 80% Hispanic<br>10% Non-Hispanic<br>10% Unknown | 9.1% Hispanic<br>90.9% Non- Hispanic | 11.1% Hispanic<br>77.8% Non- Hispanic<br>11.1% Unknown |
| **BMI (std. dev.)** | 31.9 (8.71) | 28.5 (8.08) | 27.9 (5.12) |
| **Oxygen Requirement** | 50% Room Air<br>40% Nasal Cannula<br>10% High Flow | 45.5% Room Air<br>54.5% Nasal Cannula | 100% Vented |
| **Max Temp °C (std. dev.)** | 37.3 (0.567) | 37.6 (1.01) | 38.3 (0.834) |
| **Remdesivir use** | 10% | 9.1 % | 11.1% |
| **Steroid Use** | 0% | 0% | 0% |
| **Clinical Course** | 90.0% Hospitalized<br>20% ICU | 90.9% Hospitalized<br>27.3% ICU | 100% Hospitalized/ICU |
| **Deceased** | 0% | 0% | 22.2% |
| **Length of Hospital Stay in Days (std. dev.)** | 5.0 (3.56) | 16.0 (10.7) | 35.1 (25.8) |
| **Comorbidity Score\* (std. dev.)** | 1.67 (1.29) | 2.35 (1.66) | 1.64 (1.53) |

\*Comorbidity score defined by hazard ratio for death from COVID-19 infection as determined by OPEN Safely trial (8).

### SARS-CoV-2 protein fragments identify higher levels of antigenic reactivity in older adult patients than younger adult patients

Both IgA and IgG antibodies showed higher reactivity to the full-length N protein in the older age group when compared to younger age group (Fig. 1). Within the N protein, there were significant differences with respect to age in both IgA and IgG antibody responses to protein fragments of the 151 to 419 amino acid (aa) region (Figs. 1A and 1B). Within this specified N region, protein fragments overlap to create an area of congruency in the aa 201-250 region which showed a higher IgA and IgG response in the older age group when compared to the younger age group. There was also significantly higher IgA response in the older age group compared to the younger group in the aa 351-400 region. The IgG response also differed in the N aa 351-400 region although not entirely to significance (p =0.052, fragment 301-400; p =8.2 x 10^−3^, fragment 351-419). The S1 protein, although displaying overall less reactivity than S2 and N proteins in this cohort, showed higher IgA and IgG response to aa 551-650 with overlap response at aa 551-600 in the older age group compared to the younger adult patients (Fig. 1A). Within the S2 protein, there was significant difference in IgA antibody response between older and younger patients at aa 451-550. The difference in IgG antibody response between age categories reached significance for the whole M protein, with a highlighted area of interest at the aa 1-30 region. The findings from Camerini *et al* (submitted concurrently) demonstrated similar antibody-reactive regions when looking at patients positive for SARS-CoV-2 infection compared to negative controls. Identical regions of interest occurred within the N protein at aa 151-419, S1 aa 551-650 and M aa 1-50 in this cohort and were found to have significant differences in antibody response in the older compared to younger patient groups in our cohort. Within the S2 protein, similar reactive regions were detectable in positive patients as in Camerini *et al* although these were not found to be significantly different between older and younger patients in this cohort: S2 aa 51-100, S2 aa 201-350 and S2 aa 451-480. There was, however, a significant difference detected in the IgA response to S2 aa 451-550 between age groups. While this fragment was found to be also reactive to IgG in this cohort, the difference in reactivity between age groups did not reach significance (p=0.082). The IgM response in the S2 aa 251-300 region was also found to have significant differences in reactivity between age groups. Notably, regions within the S2 protein have higher sequence homology between SARS-CoV-2 and the endemic HCoVs compared to the S1 region. In comparison, antibody response to full-length N, S1, S2 and RBD proteins by Milliplex analysis did not show significant differences between age groups (Fig. 1A) Antibody reactivity to antigenic regions were further stratified by ventilator status within the older age group, but this did not reveal any significant differences in responses when analyzed between the three groups (i.e. older ventilated, older non-ventilated and younger patients) (Fig. S1).

**Figure 1.**
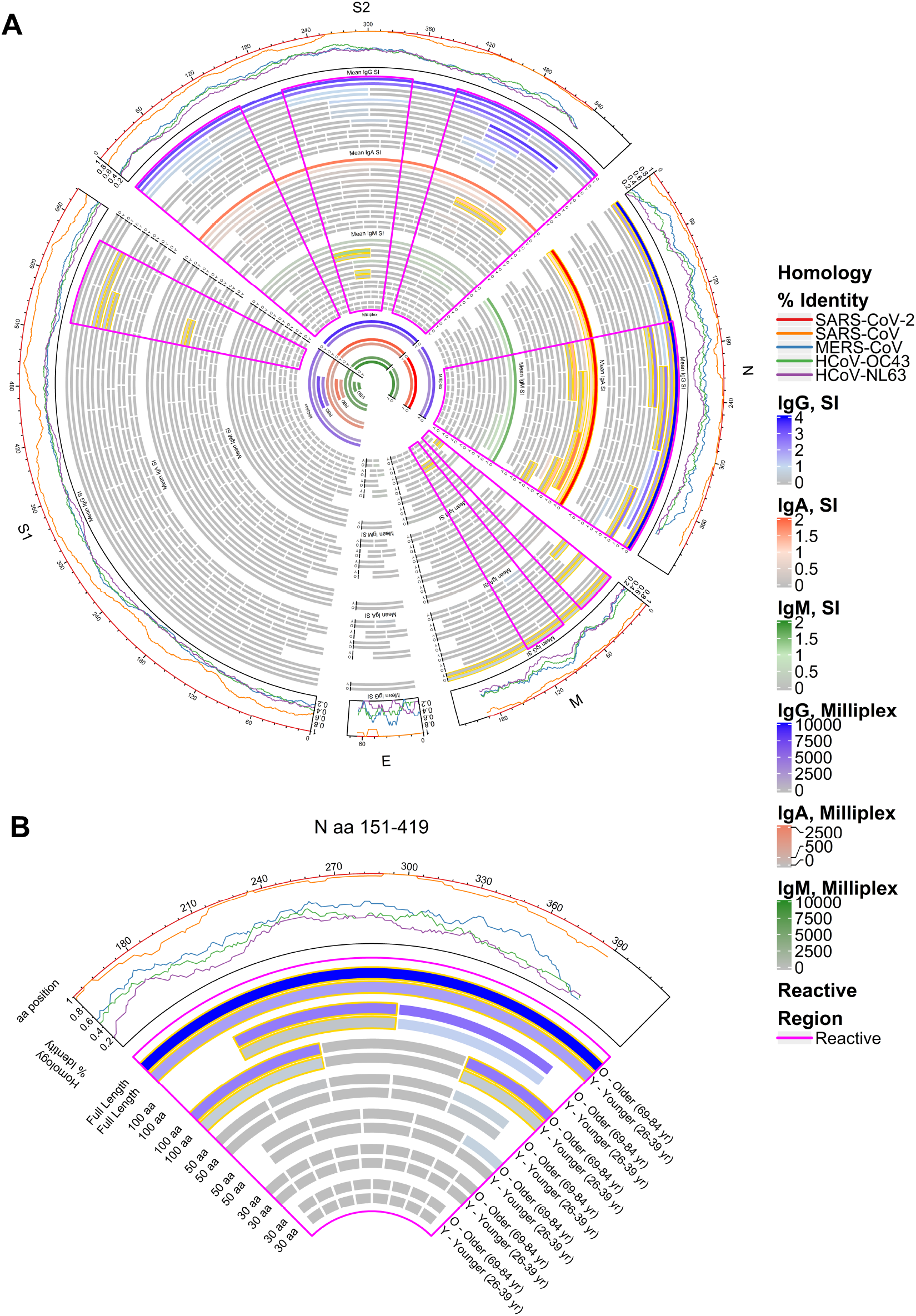
Reactivity of COVID-19 patient IgG (outer bands of bars), IgA (middle bands) and IgM (inner bands) to SARS-CoV-2 proteins displayed separately for older and younger age groups. **(A)** The circular graphic maps the amino acid (aa) position of SARS-CoV-2 fragments, showing a heat map of antibody levels in each group for overlapping regions of different aa length. Proteins are indicated outside the circle plot followed by a line graph showing the sequence homology of other HCoVs with SARS-CoV-2 for each gene. Proteins and protein fragments produced *in vitro* are indicated by bars and show length and position of each fragment in each protein. Each fragment is drawn twice and shows group mean log2 normalized signal intensity (SI) of antibody binding to each fragment for COVID-19 patient serum samples in the older age group (“O”, 69-83 yr) and the younger age group (“Y”, 26-39 yr). Signal intensity is shown by color gradients: IgG (grey to blue), IgA (grey to red), and IgM (grey to green). Bar pairs shown with gold outline represent significantly differential antibody binding between older and younger COVID-19 patients, defined as a mean log_2_ signal intensity ≥ 0.1 in at least one group and a t-test *p* value ≤ 0.05. The regions of greatest reactivity for each protein are outlined in magenta. The inner circle bands represent the responses to full-length S1, S2 and N and the receptor binding domain (RBD) in the Milliplex assay. **(B)** A sector of the circular graphic enlarged and labeled in more detail as a guide to interpreting the full figure. IgG reactivity with the C-terminal region of N protein, spanning aa sequence 151 to 419 is shown.

### Correlation of clinical data to antigenic regions of SARS-CoV-2 show association of antibodies with duration of illness

Patient serum reactive antibody responses to SARS-CoV-2 fragments were then arranged on a heat map for comparison to patient clinical characteristics and responses to other HCoVs (Fig. 2A). This demonstrated overall higher reactivity of antibody response among the older patients compared to the younger patients. This was most striking in IgG responses but could also be seen in IgA responses, although with varying consistency in reactivity levels between the two isotypes (i.e. a patient with high IgG levels to a specific SARS-CoV-2 fragment did not always have corresponding high levels of reactivity with IgA and visa-versa). However, antibody reactivity displayed substantial heterogeneity within age and severity groups with some patients showing little to no IgG, IgA and IgM response to all SARS-CoV-2 fragments. Notably, patients without antibody reactivity to protein fragments did have reactivity to proteins of other HCoVs. The serum IgG response to HCoV-OC43 and HCoV-NL63 appeared robust in most patients, but serum IgA and IgM responses to these HCoVs had less signal intensity.

**Figure 2.**
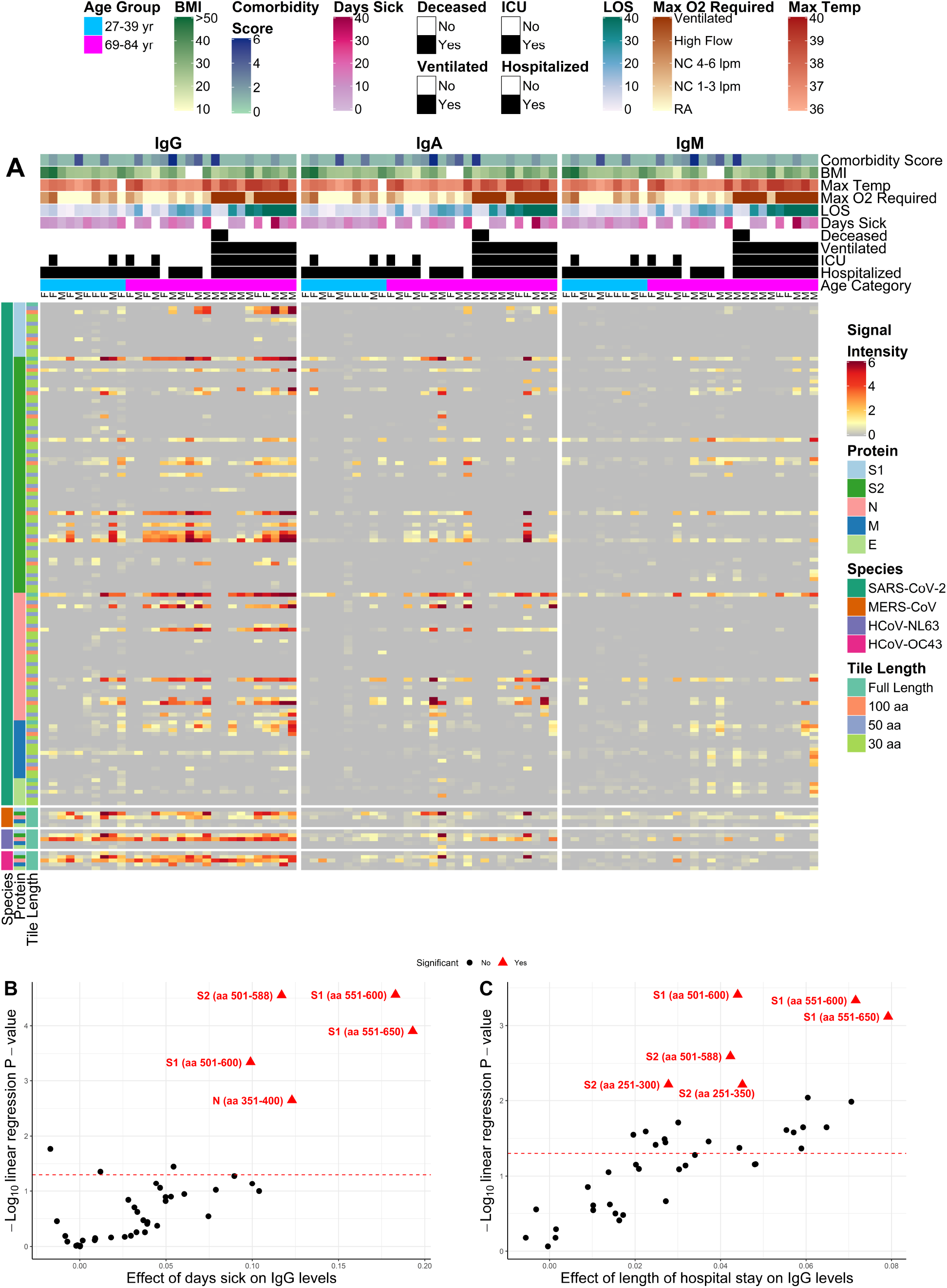
Heatmap depicting relative IgG (left panel), IgA (middle panel) and IgM (right panel) antibody responses to SARS-CoV-2 as compared to other HCoVs and clinical data. **(A)** The heatmaps present the signals of antibody binding to individual proteins and protein fragments within the antigenic regions of SARS-CoV-2, as well as the full-length structural proteins of MERS-CoV, HCoV-NL63 and HCoV-OC43, for individual samples. Columns represent serum samples, and rows represent proteins or protein fragments; 128 SARS-CoV-2 proteins or fragments, five proteins each of MERS-CoV, HCoV-OC43 and HCoV-NL63. Antibody log_2_ signal intensity is shown on a color scale from grey to red. Sample clinical information is overlaid above the heatmaps and includes sex (M/F), age category, clinical status (hospitalized, admitted to ICU, ventilated and/or deceased), longevity of symptoms (days sick prior to sample collection and length of stay “LOS” at hospital), maximum oxygen levels required and patient measurements including maximum body temperature, body mass index (“BMI”) and composite score encompassing patient’s other comorbidities (“Comorbidity Score”). Protein/fragment information is annotated to the left of the heatmaps and includes the virus, full-length protein name and the amino acid length of the protein fragments (“Tile Length”, as full length, 100, 50 or 30 aa). **(B-C)** The volcano plots show the statistical effect estimates of days sick prior to serum sample collection and the length of hospital stay on IgG levels, respectively. The x-axis shows the linear regression coefficients that were adjusted by age category, sex and requirement of ventilator, and the y-axis shows the inverse log_10_ p values for each of the SARS-CoV-2 proteins that were reactive (normalized log_2_ signal intensity > 1.0) in at least 10% of the study population. The proteins/fragments showing significant associations after correction for the false discovery rate are highlighted as red triangles and red labels.

Linear models were created to observe correlations between patient clinical data and antibody binding to SARS-CoV-2 fragments which were reactive in at least 10% of the population. After adjustment for age, sex and ventilator status, a significant correlation was found between IgG response and days from symptom onset (Fig. 2B). A region of notable correlation was found in the S1 aa 551-600, which was further supported by significant correlation seen with S1 aa 551-650 fragment and S1 aa 501-600 fragment. S2 aa 501-588 was additionally found to have significant correlation with days from symptom onset. There was also a significant correlation found between IgG antibody response and length of hospital stay (Fig. 2C). As with days of illness, this correlation was found to be most notable regarding the S1 aa 551-600 region, further supported by significant correlation in the S1 aa 551-650 and S1 aa 501-600 fragments. Additional correlation with length of hospital stay was also seen in the S2 aa 501-588 region.

### Serum cytokine and chemokine profiles correlate with antibodies in older adult COVID-19 patients more than in younger adult patients

We then assessed the association between antibody reactivity to SARS-CoV-2 fragments adjusted for age, sex and ventilator status, and cytokine levels in each patient sample analyzed with the MILLIPLEX® MAP Human Cytokine/Chemokine/Growth Factor Panel (48 Plex; Fig S2). There were significantly positive correlations seen in IgG, IgM and IgA antibody response and levels of interleukin-5 (IL-5), tumor necrosis factor-β (TNF-β), platelet derived growth factor-AB/BB (PDGF-AB/BB), epidermal growth factor (EGF), soluble CD40 ligand (sCD40L) and interleukin-17A (IL-17A) in serum samples. Negative correlations were seen between antibody responses and levels of interleukin-10 (IL-10), interferon gamma-induced protein 10 (IP-10, or chemokine ligand 10, CXCL10), interferon-α2 (IFN-α2), tumor necrosis factor-α (TNF-α) and interleukin-15 (IL-15).

Correlations between antibody reactivity to antigenic protein fragments and cytokine/chemokine levels in patient serum samples were then stratified by age group (Fig. 3). This revealed the same positive and negative correlations consistently represented in the older age group. In contrast, the younger adult patient group demonstrated notable heterogeneity in its correlations with antibody responses. Furthermore, correlations that were significantly positive or negative in the older age group at times showed a reverse correlation in the younger age group. For instance, IL-10 was significantly negatively correlated to IgG response most notably to the N and S2 fragments in the older age group. This, however, was not consistent with the younger age group where correlations between IgG response to these regions and IL-10 are, although variable, mostly positive. However, similar correlations did occur regardless of age group, such as with the significantly positive correlation seen between the IgG response to N aa 200-400 and levels of IL-5.

**Figure 3.**
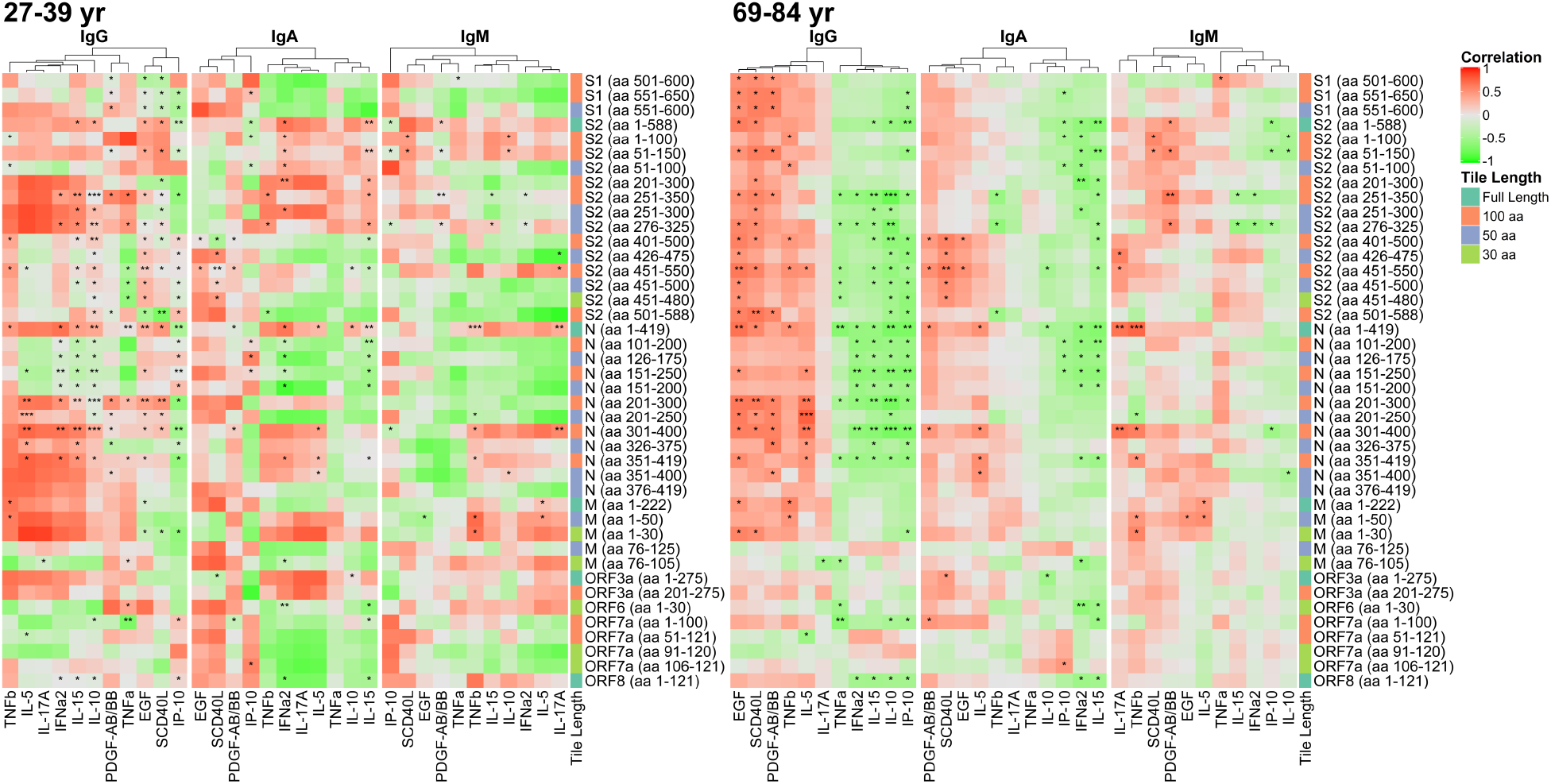
Correlation between reactive SARS-CoV-2 proteins and fragments with selected cytokine/chemokine levels stratified by age group. The heatmap shows the Pearson’s correlation coefficient between antibody and cytokine levels on a colorimetric scale from negative correlation in green to positive correlation in red. Significance of the correlations are shown by overlaid asterisks (^*^ = p<0.05, ^**^ = p<0.005, ^***^ = p<0.0005). Plots are separated by the younger age group (26-39 yr, left) and older age group (69-83 yr, right). The antigens displayed correspond to proteins and fragments produced *in vitro* that were seropositive (≥ 1.0 log_2_ normalized signal intensity) in at least 10% of the study population. The cytokines displayed were selected based on significant associations with antibody levels in linear mixed effects regression models, adjusted for age category, sex and requirement of ventilator. IL-17A and IL-5 were selected due to significant associations with individual antibody responses in ordinary least squares regression models adjusted for age category, sex and ventilator. Protein/fragment information is annotated to the right of the heatmaps and includes the protein name and the amino acid coordinates in parentheses and the length of the protein fragments (“Tile Length”).

### IgG responses to SARS-CoV-2 S2 protein correlate with IgG responses to homologous endemic human coronavirus S2 proteins

We further assessed the correlation of the IgG response to the S2 and N proteins of SARS-CoV-2 to that of HCoV-OC43 and HCoV-NL63 (Fig. 4). There were strong linear correlations seen between antibody reactivity to the S2 protein of SARS-CoV-2 and the S2 proteins of HCoV-OC43 and HCoV-NL63 regardless of age or ventilator status (Figs. 4, S3A and S3B). This was inconsistent with correlations observed between IgG responses to N proteins, which were weakly correlated, in part due to individuals with little or no reactivity (log_2_ normalized signal intensity < 1.0), that suggested a population with either delayed or negative seroreactivity, henceforth “serosilent” individuals. This was further exemplified through density plots highlighting the differences in bimodal antibody responses between responding and non-responding individuals. Notably these individuals were responsive to S2 and N proteins of endemic HCoVs and appeared to be non-responsive to solely SARS-CoV-2 proteins (Fig. 4).

**Figure 4.**
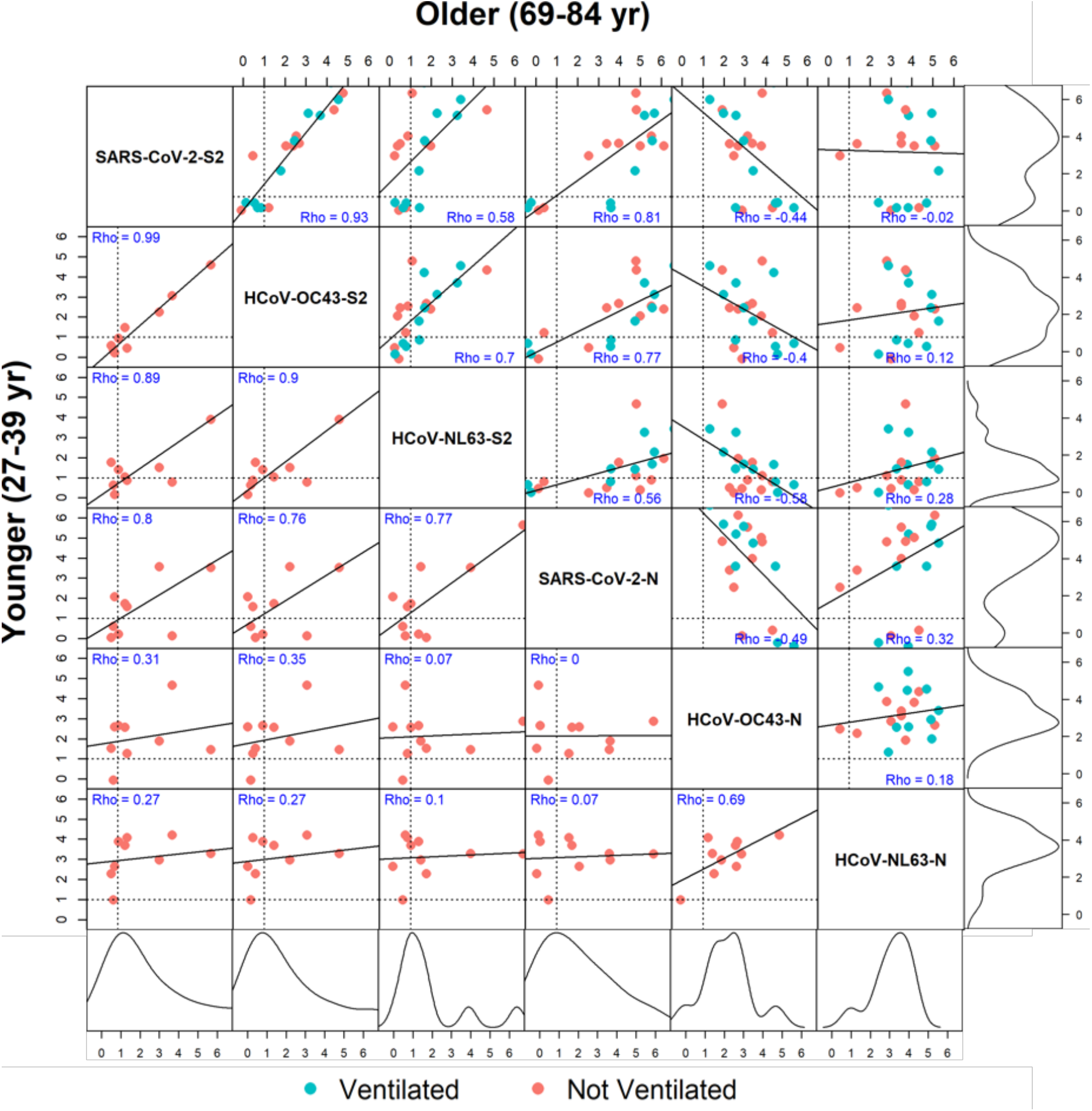
Correlation of IgG response to full-length proteins of SARS-CoV-2 and two endemic human coronaviruses. Correlogram depicting the Spearman’s rank correlation coefficient (*Rho*) between IgG normalized signal intensity to SARS-CoV-2, HCoV-OC43 and HCoV-NL63 full-length S2 and N proteins produced *in vitro*. The lower left half of the diagonal shows correlations between reactivity of sera in the younger age group (27-39 yr), and the upper right half of the diagonal shows the older group (69-84 yr). Ventilated patients are represented by teal dots and non-ventilated patients are represented by red dots. Lines of seropositivity defined as a normalized log_2_ signal intensity ≥ 1 are depicted by horizonal and vertical dotted lines within each scatterplot. The *Rho* coefficient is listed in blue lettering in each box. Outer most right and bottom boxes represent density plots for older and younger age groups, respectively.

Positive correlations between S2 responses to SARS-CoV-2 and endemic HCoVs were not observed for IgM, largely owing to the limited IgM response seen to HCoV-OC43 and HCoV-NL63 as discussed earlier (Fig. S4A). While similar correlations as with IgG were seen when comparing IgA responses to S2 proteins of endemic HCoVs, this did not extend to N proteins, again, due to the limited IgA response in serum (Fig. S4B). When comparing the antibody response to individual SARS-CoV-2 fragments to endemic coronaviruses, there were diffusely positive correlations seen between S2 fragments with S2 proteins of endemic coronaviruses, most notably to HCOV-OC43 (Fig. S5).

Given the notable separation seen in non-responding individuals compared to the rest of the cohort, cytokine profiles were then assessed in these patients. As the majority of these individuals were in the older age group, cytokines/chemokines from the older age group were compared (Fig. 5). We identified four serosilent individuals to both immunodominant SARS-CoV-2 proteins, N and S2 (N-, S2-), with an additional two patients that were seronegative to S2 protein (S2-) but seroreactive to N protein (N+) (Fig. S6). Analysis of cytokine/chemokine responses to all six patients revealed significantly higher IL-10, IL-15 and IP-10 in serosilent individuals when compared to the rest of the older age group. It also displayed significantly lower levels of EGF and sCD40L in serosilent individuals (Fig. 5).

**Figure 5.**
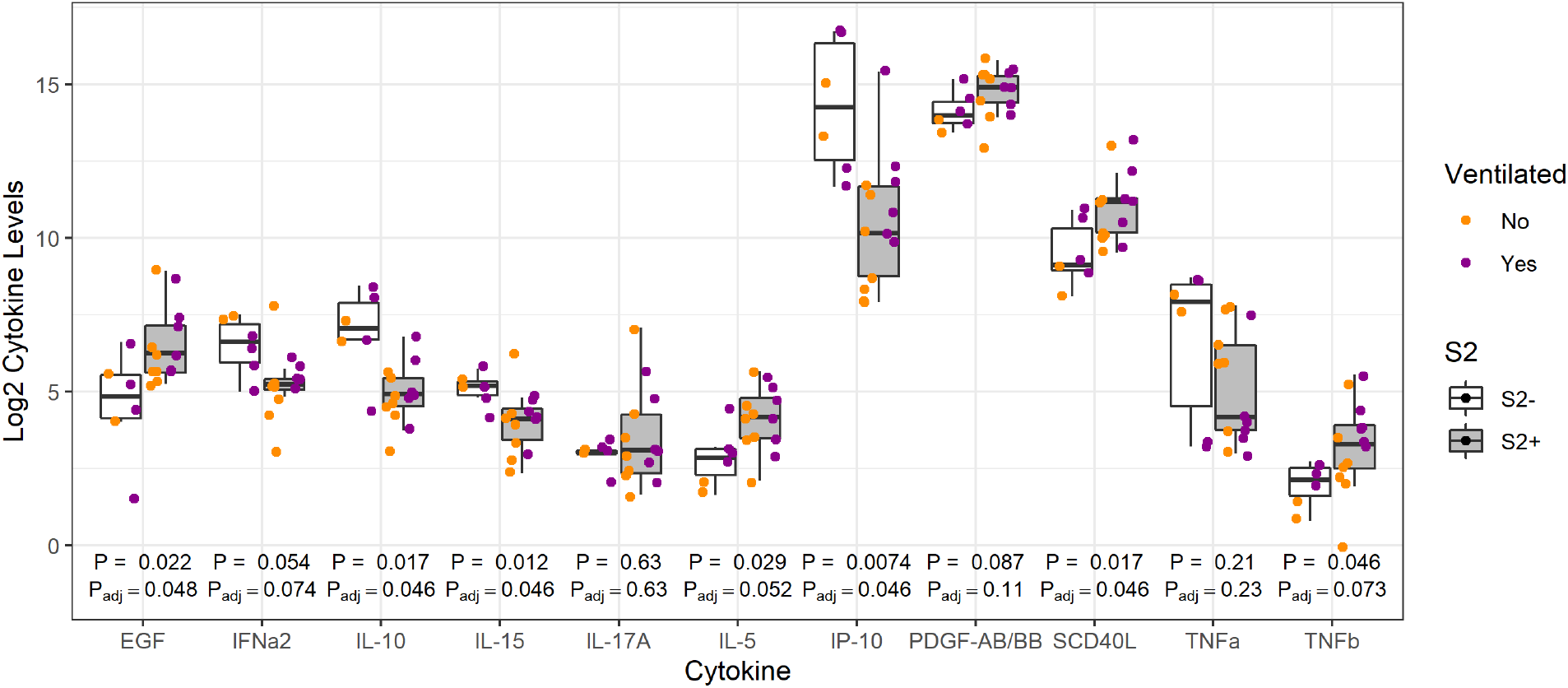
Differences in cytokine and chemokine levels between serosilent and seroreactive older adult COVID-19 patients. The boxplot illustrates the top eleven differences in cytokine levels of those nonresponding to SARS-CoV-2 full length S2 proteins (S2-“serosilent”; represented by white boxes) and those responding (S2+ “seroreactive”; represented by grey boxes). Cytokine/chemokine levels are plotted on a log_2_ scale. Unadjusted Wilcoxon’s rank sum p-values are denoted underneath each pair of boxes with an asterisk above p-values that remain significant after correction for the false discovery rate.Ventilated patients are represented by blue dots and non-ventilated patients are represented by orange dots.

## Discussion

While this study revealed epitopes seen within SARS-CoV-2 proteins, which corresponded to the concurrent study by Camerini *et al*, it additionally revealed how reactivity differed between age groups and how these same regions correlated with clinical and laboratory data. Although differences between antibody responses did not reach significance when separated by severity (i.e., those ventilated vs those who did not require ventilation), there were significant differences seen between age groups, with higher antibody reactivity apparent in the older age group. We were additionally able to see correlations in antibody response to SARS-CoV-2 antigenic fragments and serum cytokine/chemokine profiles along with correlations in reactivity to endemic HCoVs.

Among the regions of interest highlighted in our study was S1 aa 551-600 region, an area just past the RBD at the C-terminus of the S1 protein, which has also been noted in other recent studies involving SARS-CoV-2 epitope mapping (12,13). In our study, IgG reactivity to this region was not only higher in the older COVID-19 patient cohort but was significantly correlated with hospital length of stay and days of illness. While differences in reactivity did not reach significance regarding ventilator status in this small cohort, the correlations found in this study suggest unfavorable clinical outcomes associated with large levels of antibodies to this region.

Previous studies have also shown a correlation between IgG responses to S1 protein and days of illness, which likely can be attributed, at least in some part, to reactivity within this region (14).

We further highlighted three regions within the S2 protein: S2 aa 51-100, S2 aa 201-350 and S2 aa 451-480, which have also been implicated in recent epitope mapping studies (12,13). While small sample size may have limited our ability to see true differences in IgG reactivity within these regions with respect to severity and age, we were able to see a correlation between reactivity within the S2 aa 201-350 and hospital length of stay, highlighting clinical implications to having antibodies to this region.

When antibody reactivity to SARS-CoV-2 was correlated with cytokine/chemokine profiles in these serum samples, we found consistent correlations existed within the older patient group while large variations occurred in younger COVID-19 patients. This suggests a clinically unfavorable cytokine/chemokine profile that correlates with higher antibody reactivity, which more commonly occurs in older patients. IL-5, a type 2 (Th_2_) cytokine shown by Lucas *et al* to correlate with severe COVID-19 disease, was shown to have a significantly positive correlation to antibody response to the N aa 200-400 region, which may further suggest poor outcomes related to Th_2_ responses (15). Additionally, IL-10 had significant negative correlations to antibody responses to S2 and N proteins in the older age group, which is consistent with its known anti-inflammatory properties. Interestingly IL-10 has been implicated in numerous other viral, bacterial, and protozoal infections whose clinical outcomes were observed to be time-dependent of peak IL-10 production and its ability to cause either inhibition of effective pathogen clearance or prevention of excessive immune response to foreign infectious antigens (16).

We also found strong correlations between the IgG response to SARS-CoV-2, HCoV-NL63 and HCoV-OC43 S2 proteins, which have also been noted in other recent epitope studies (12,13). This has been attributed to considerable sequence homology observed between S2 proteins of SARS-CoV-2 and endemic HCoVs, particularly to the more closely related endemic betacoronaviruses (HCoV-OC43 and HCoV-HKU1). Associations found in this study suggest either cross-reactivity of newly produced antibodies to SARS-CoV-2 with other HCoV antigens in the array or cross-reactivity in which preexisting antibodies to other coronaviruses can recognize SARS-CoV-2 antigens. While this is difficult to determine without analysis of patient serum prior to infection, we see evidence of both phenomena occurring in our cohort. Lack of a concomitant serum IgM response observed in this cohort to endemic HCoVs along with lack of observed correlation between the IgM response to S2 proteins between them suggests a preexisting, boosted IgG rather than new, acute antibody production. However, the magnitude of reactivity to S2 protein and the positive correlation of anti-S2 IgG between SARS-CoV-2 and endemic HCoVs suggests likely a component of new antibody reactivity to some epitopes due to significant immune activation. Preexisting, cross-reacting antibodies to SARS-CoV-2 would allow an opportunity for cross neutralization of SARS-CoV-2 antigens and raises the possibility of improved clinical outcomes in these patients. This may further explain why children, who are known to have more consistent exposures to endemic HCoVs, may be more protected from severe COVID-19 infection (12). However, of note, in our analysis, correlations of antibodies to S2 proteins between SARS-CoV-2 and endemic HCoVs were apparent regardless of age or ventilator status, possibly suggesting less of an influence on clinical outcomes.

There was also notably absent antibody reactivity to SARS-CoV-2 proteins among a subset of individuals in this cohort. Given that these patient samples were collected at a single time point, it is difficult to know if these represent patients with no response throughout the entire illness course or are individuals in which antibody levels were slow to respond. Wajnberg *et al* found the latter in assessment of longitudinal samples, noting that in addition to a slow antibody response to SARS-CoV-2 infection, peak titers were lower than patients with a more robust initial response (17). This may be clinically relevant, as patients with low and slow antibody response may be those likely to benefit most from SARS-CoV-2 antibody treatment regimens.

We then further characterized serosilent patients by looking at their clinical characteristics and immune response. Although conclusive analysis is limited by sample size, two out of the four serosilent (N-, S2-) older adults required ventilation, three were admitted to the ICU and two were the only deceased patients in the study, suggesting negative clinical outcomes associated with minimal antibody response in these patients. In contrast to severe COVID-19 disease linked with high antibody response as discussed above, serosilent patients suggest an alternative immunologic profile to infection providing an additional avenue for poor clinical outcomes. Among the cytokine differences discovered between serosilent and responsive patients, IL-10 was implicated as one of the most differential, with serosilent individuals displaying significantly higher levels compared to the rest of the older patient cohort. This is again congruent with known influences of IL-10 as discussed above and highlights its potential role in the serosilent patient cytokine profile (16). We additionally found a significant decrease in sCD40L in serosilent patients compared to the rest of the group, which is consistent with sCD40L’s known ability to promote B cell proliferation, differentiation and immunoglobulin production (18).

A substantial limitation in our study was the small sample size which likely limited our ability to detect relationships between epitopes, cytokines, and clinical outcomes. This further limited in our ability to statistically analyze and classify our serosilent samples and therefore were categorized subjectively based on full length IVTT N and S2 protein reactivity. As this small cohort is meant for hypothesis generation, a larger cohort is needed to further validate our findings. Our study is also limited to epitopes produced in *Escherichia coli* which restricts our ability to see epitopes that require eukaryotic post-translational modification such as glycosylation. This is particularly relevant in regards to the spike protein, which exists as a trimer on the virion surface and undergoes conformational changes during viral entry into cells (6). As addressed in Camerini *et al* we also observed similar limitations in response to S1 fragments produced *in vitro*, which perhaps was influenced in part by prokaryotic production of IVTT proteins. However, we were able to detect an area in S1 which is notable in its correlations and outcomes as discussed above.

## Methods

### Patient sample and clinical data collection

Patients who tested positive for COVID-19 by PCR at the University of Virginia Medical Center had residual routine lab specimens collected into a biorepository. Serum samples of patients of varying age and severity were collected from April 2020 until July 2020. Blood collected in EDTA was centrifuged at 1300 x g for 10 minutes, then plasma was aliquoted and stored at − 80°C until testing. Thirty of these serum samples were provided to Antigen Discovery Inc. to be exposed to protein microarrays as described below.

Clinical data including patient medical history, lab work and clinical course were collected from the electronic medical record using honest brokers with unique study numbers to ensure confidentiality (Tables 1, S2). Days from symptom onset were determined through assistance from an honest broker who read through history and physical exam notes, emergency department notes, progress notes and discharge summaries of patients with COVID-19. Comorbidity scores were derived from hazard ratios presented in the OPEN Safely trial by Williamson *et al* to appropriately weigh patient comorbidities with previously observed associations in risk of death from COVID-19 (8). Collection of blood samples and deidentified patient information was approved by the University of Virginia Institutional Review Board IRB-HSR #22231 and 200110.

### Protein microarray analysis of serum samples

The first generation multi-coronavirus protein microarray, produced by Antigen Discovery, Inc. (ADI, Irvine, CA, USA), included 935 full-length coronavirus proteins, overlapping 100, 50 and 30 aa protein fragments and overlapping 13-20 aa peptides from SARS-CoV-2 (WA-1), SARS-CoV, MERS-CoV, HCoV-NL63 and HCoV-OC43. Purified proteins and peptides were obtained from BEI Resources. All these coronavirus proteins were produced in *Escherichia coli* except the SARS-CoV-2 and SARS-CoV S proteins, which were made in Sf9 insect cells and the SARS-CoV-2 RBD, made in HEK-293 cells. Other proteins and protein fragments were expressed using an *E. coli in vitro* transcription and translation (IVTT) system (Rapid Translation System, Biotechrabbit, Berlin, Germany) and printed onto nitrocellulose-coated glass AVID slides (Grace Bio-Labs, Inc., Bend, OR, USA) using an Omni Grid Accent robotic microarray printer (Digilabs, Inc., Marlborough, MA, USA). Microarrays were probed with sera and antibody binding detected by incubation with fluorochrome-conjugated goat anti-human IgG or IgA or IgM (Jackson ImmunoResearch, West Grove, PA, USA or Bethyl Laboratories, Inc., Montgomery, TX, USA). Slides were scanned on a GenePix 4300A High-Resolution Microarray Scanner (Molecular Devices, Sunnyvale, CA, USA), and raw spot and local background fluorescence intensities, spot annotations and sample phenotypes were imported and merged in R (R Core Team, 2017), in which all subsequent procedures were performed. Foreground spot intensities were adjusted by subtraction of local background, and negative values were converted to one. All foreground values were transformed using the base two logarithm. The dataset was normalized to remove systematic effects by subtracting the median signal intensity of the IVTT controls for each sample. With the normalized data, a value of 0.0 means that the intensity is no different than the background, and a value of 1.0 indicates a doubling with respect to background. For full-length purified recombinant proteins and peptide libraries, the raw signal intensity data was transformed using the base two logarithm for analysis.

### Milliplex serum analysis

Data from the protein microarray was compared to the same thirty samples analyzed with MILLIPLEX ® SARS-CoV-2 Antigen Panel 1 IgG, IgA, and IgM for comparison (Millipore Sigma, St. Louis, MO). IgG, IgA and IgM antibodies are captured by specific bead-region microspheres, each conjugated with SARS-CoV-2 antigen subunits S1, S2, RBD, or N, and are measured by median fluorescent intensity (MFI). The four antigens are recombinant poly-his-tagged. Kit instructions were followed. Samples were diluted 1:100 in assay buffer. Ninety-six well plates were pre-wetted with 200 μL wash buffer, covered with plate sealer and incubated for 10 minutes at room temperature with shaking, then emptied. To all wells, 25 μL of Assay Buffer was added. Twenty-five μL of each diluted sample was added to the sample wells and 25μL of Assay Buffer was added to background wells. Sixty μL of both sonicated (30 seconds) and vortexed (1 minute) analyte and control bead was combined and brought to a final volume of 3 mL with the addition of Assay Buffer. After vortex, 25 μL of bead mixture was dispensed into each plate well. The plate was sealed and incubated for 2 hours at room temperature with constant shaking. A handheld magnetic plate washer was used to retain magnetic beads while liquid contents were discarded appropriately. Wells were washed 3 times with 200 μL wash buffer. Fifty μL of phycoerythrin-anti-human immunoglobulin (IgG, IgA or IgM per kit in use) detection antibody was added to each well, plate sealed and incubated 90 minutes at room temperature with constant shaking. Plates were washed three more times with magnetic plate washer. 150 μL sheath fluid was added to each well, the plate was then sealed and shaken at room temperature for 5 minutes. The plate was then read on a Luminex ® MAGPIX™ Instrument System with a minimum of 50 beads of each analyte collected per well. Cytokine levels were additionally measured in each sample via MILLIPLEX® MAP Human Cytokine/Chemokine/Growth Factor Panel A (48-Plex) through similar methods.

### Statistical Analysis

Student’s t-tests were used for comparison of the individual antibody response means between the younger and older age groups. Proteins or protein fragments expressed using the IVTT system were classified as reactive antigens based on a 1.0 normalized signal intensity seropositivity threshold and seroprevalence cutoff of 10% of the study population (i.e. at least 3 seropositive responses) for IgG, IgA or IgM. Multivariable ordinary least squares (OLS) regression was used to model associations between antibody and patient information obtained from electronic records. Antibody responses to individual reactive antigens (n=52) were modeled as dependent variables, and the following variables were modeled as independent variables: sex, age category, requirement of a ventilator, days symptomatic prior to sample collection, length of hospital stay, admission to the ICU, maximum required supplemental oxygen category, comorbidity score, maximum body temperature during while admitted, body-mass index (BMI), maximum CRP, maximum ferritin, maximum D dimer, minimum lymphocytes, maximum AST and troponin lab levels, and the base 2 log-transformed measurements from the Milliplex serum analysis. Due to the moderate sample size of the study, not all independent variables were modeled simultaneously. Three “base” variables were used to adjust the effect estimates of all other independent variables in separate 4-variable models; these base variables were sex, age category and requirement of a ventilator. Adjustment for the false discovery rate was performed using the “p.adjust” function in R (19). To select variables associated with SARS-CoV-2-specific antibodies, linear mixed effects regression (LMER) was used to model all antibody responses against SARS-CoV-2 reactive antigens with random intercepts at the sample level and antigen level to adjust for repeated measures. Similar to the approach with OLS regression, LMER models used the same 3 base variables to fit separate models for all other fixed effects variables. All coefficients were returned from models fit using restricted maximum likelihood (REML). To generate P-values for LMER models, the models were refit using maximum likelihood (ML) and compared by ANOVA against null models with the coefficient removed using ML. Cytokines and chemokines that were significantly associated with antibody levels in LMER models were correlated with SARS-CoV-2 reactive antigens using Pearson’s correlation coefficient. Clinical patient variables were associated with cytokine levels using OLS regression, similarly to antibody models. Correlation between SARS-CoV-2 S2 and N proteins with HCoV-OC43 and HCoV-NL63 S2 and N proteins was assessed using Pearson’s correlation coefficient. Samples were categorized as “serosilent” if full-length S2 IgG responses were less than 1.0 normalized signal intensity. Differences in median log_2_ cytokine levels between serosilent and seroreactive subjects was assessed using Wilcoxon’s rank sum test. Data visualization was performed using the circlize (20), ComplexHeatmap (21), ggplot2 and corrplot (22)packages in R. The p-values presented for full-length and overlapping fragments of SARS-CoV-2 proteins were not adjusted for the false discovery rate, because the measurements are not independent and an appropriate method of p-value correction was not to our knowledge available for the extent of dependence in the antibody measurements. As expected, there were high levels of colinearity in the antibody response to overlapping fragments of different sizes in the reactive regions of SARS-CoV-2 proteins. Although unadjusted p-values were used in these comparisons, the concordance of fragment antibody binding and differential immunoreactivity in the independent study reported concurrently in Camerini *et al* lends confidence that the responses reported are unlikely to be due to chance. The full results from linear models are included in Supplemental File 1. However, further studies will be able to validate these findings.

## Data Availability

All data will be made available.

