## Supplement for "Diverse Humoral Immune Responses in Younger and Older Adult COVID-19 Patients"

[1]Department of Medicine, University of Virginia Health System, Charlottesville, VA, USA; [2]Antigen Discovery Incorporated (ADI), Irvine CA, USA; [3]Department of Microbiology, Immunology and Cancer Biology, University of Virginia Health System, Charlottesville, VA, USA; [4]Department of Pathology, University of Virginia Health System, Charlottesville, VA, USA; [5]Center for Virus Research, University of California, Irvine, CA, USA

### Supplement

**Supplemental Table S1. Features of the 1st Generation ADI Multi-Coronavirus Protein Microarray**

| Virus | S-protein | E-protein | M-protein | N-protein | Accessory or Other Proteins | Total Number of Features = 1007 |
|---|---|---|---|---|---|---|
| SARS-CoV-2 | RBD*, S# from BEI, S1, S2, 30, 50 & 100 aa fragments by IVTT at ADI | Protein and 30, 50 aa fragments by IVTT at ADI | Protein and 30, 50, 100 aa fragments by IVTT at ADI | Protein BEI, 30, 50 & 100 aa fragments by IVTT at ADI | ORF 3a, 6, 7a, 8 and 10 protein, 30, 50 & 100 aa fragments by IVTT at ADI | 11 proteins, 311 fragments = 322 features |
| SARS-CoV | Protein** and peptide set from BEI Resources | IVTT by ADI | Protein, peptides from BEI | Protein, peptides from BEI | 3CL protease | 5 proteins, 208 peptides = 213 features |
| MERS-CoV | BEI Resources | IVTT by ADI | IVTT by ADI | BEI Resources | ORF 3a, 4a, 4b, 5 and 8b protein IVTT by ADI | 9 proteins |
| HCoV-NL63 | Protein by IVTT at ADI, peptides from BEI | IVTT by ADI | IVTT by ADI | IVTT by ADI | ORF 3 protein IVTT by ADI | 5 proteins and 226 peptides = 231 features |
| HCoV-OC43 | Protein by IVTT at ADI, peptides from BEI | IVTT by ADI | IVTT by ADI | IVTT by ADI | HE, N2 protein IVTT by ADI | 6 proteins and 226 peptides = 232 features |

IVTT means coupled *in vitro* transcription and translation. * RBD is the receptor binding domain, aa 319 to 541 of the SARS-CoV-2 S protein. # SARS-CoV-2 S protein is a stabilized form with a trimerization sequence and transmembrane domain deletion. ** SARS-CoV-S protein is a transmembrane domain deleted form.

**Supplemental Table S2. Median and interquartile range of maximum lab values for admission of the young adult, older non ventilated, and older ventilated COVID-19 patients.**

| | **Young Adult Patients (N=10; Age 27-39)** | **Older Patients, Not Ventilated (N=11; Age 69-82)** | **Older Patients, Ventilated (N=9; Age 69-83)** |
|---|---|---|---|
| **C-Reactive Protein, mg/dL (interquartile range)** | 7.70 (0.9-12.7) | 7.85 (5.15-13.3) | 9.80 (1.8-24.1) |
| **Ferritin, ng/mL (interquartile range )** | 188 (64-925) | 704 (407-993) | 2596 (1238-4003) |
| **D dimer, ng/mL (interquartile range)** | 345 (203-461) | 219 (169-580) | 1882 (1049-2356) |
| **Minimum Lymphocyte Count, K/ul (interquartile range)** | 1.02 (0.52-1.07) | 0.90 (0.840-1.32) | 0.620 (0.43-1.44) |
| **Aspartate Aminotransferase, U/L (interquartile range)** | 60.0 (43.5-98.5) | 42.5 (36-65) | 74.0 (70-114) |
| **Troponin, ng/mL (interquartile range)** | 0.030 (-) | 0.030 (0.03-0.08) | 0.040 (0.03-0.17) |

**Supplemental Figure S1**

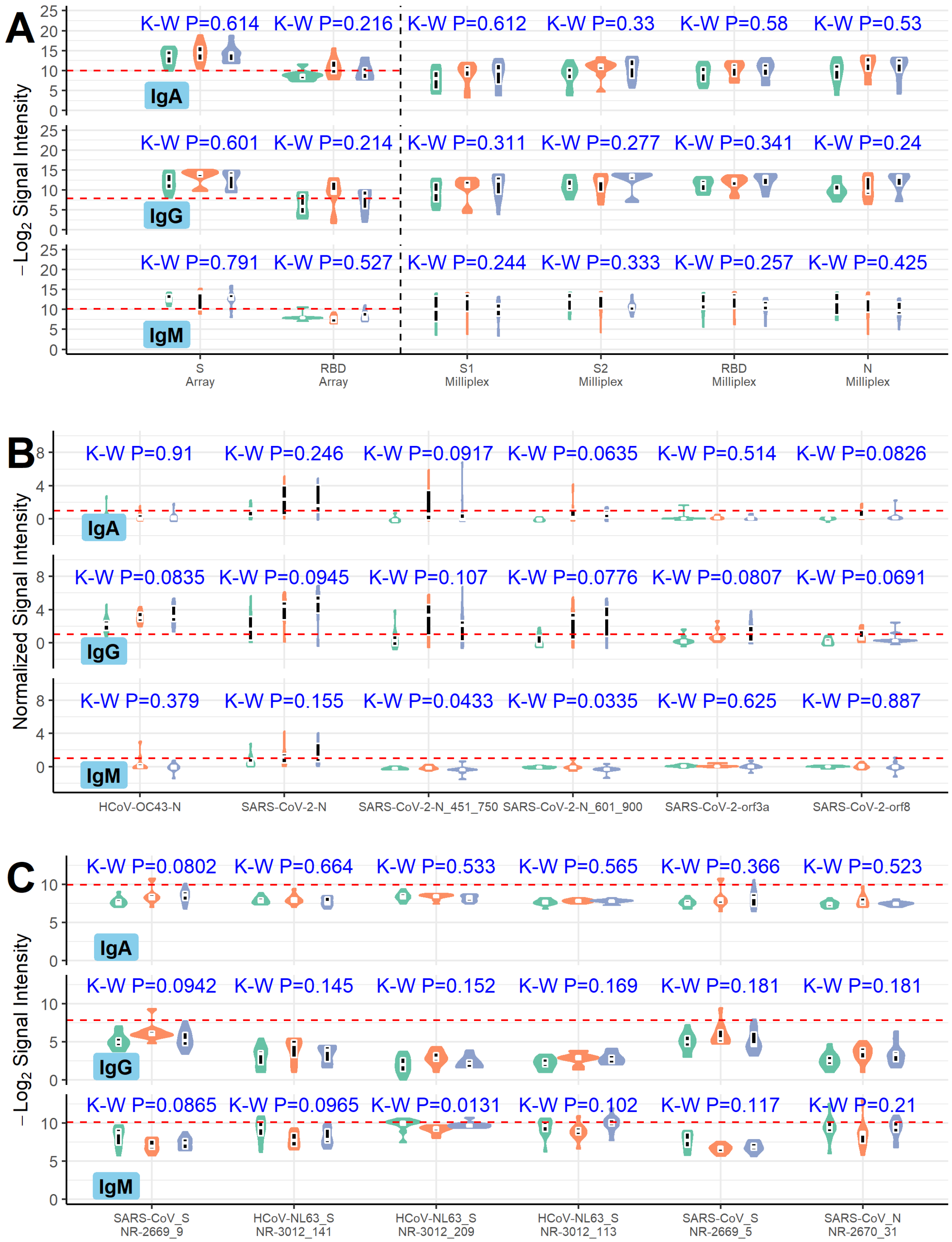

**Supplemental Figure S1: Differential analysis of serum antibody responses by age category and requirement of ventilator. (A-C)** The violin plots show the distribution of signals to selected proteins and fragments. Within each violin is a nested trimmed boxplot in black showing the interquartile ranges, and above each set of violins is the unadjusted p-value from Kruskal-Wallis 3-group tests. **(A)** Responses to recombinant purified S and RBD proteins printed the multi-coronavirus microarray (left of vertical black dashed line) are shown in comparison with responses to recombinant purified S1, S2, RBD and N proteins used in the Milliplex assay (right of the vertical black dashed line). The horizontal red dashed is a threshold representing the global mean and 3 standard deviations of all purified proteins and peptides on the array. **(B)** The six proteins or fragments produced *in vitro* with the greatest differential reactivity between the three groups are shown with a horizontal red dashed line representing the seropositivity threshold of 1.0 $\log_2$ normalized signal intensity. **(C)** The six peptide library targets with the greatest differential reactivity between the three groups are shown with a horizontal red dashed line representing the mean and 3 standard deviations of all purified proteins and peptides on the array.

#### Supplemental Figure S2

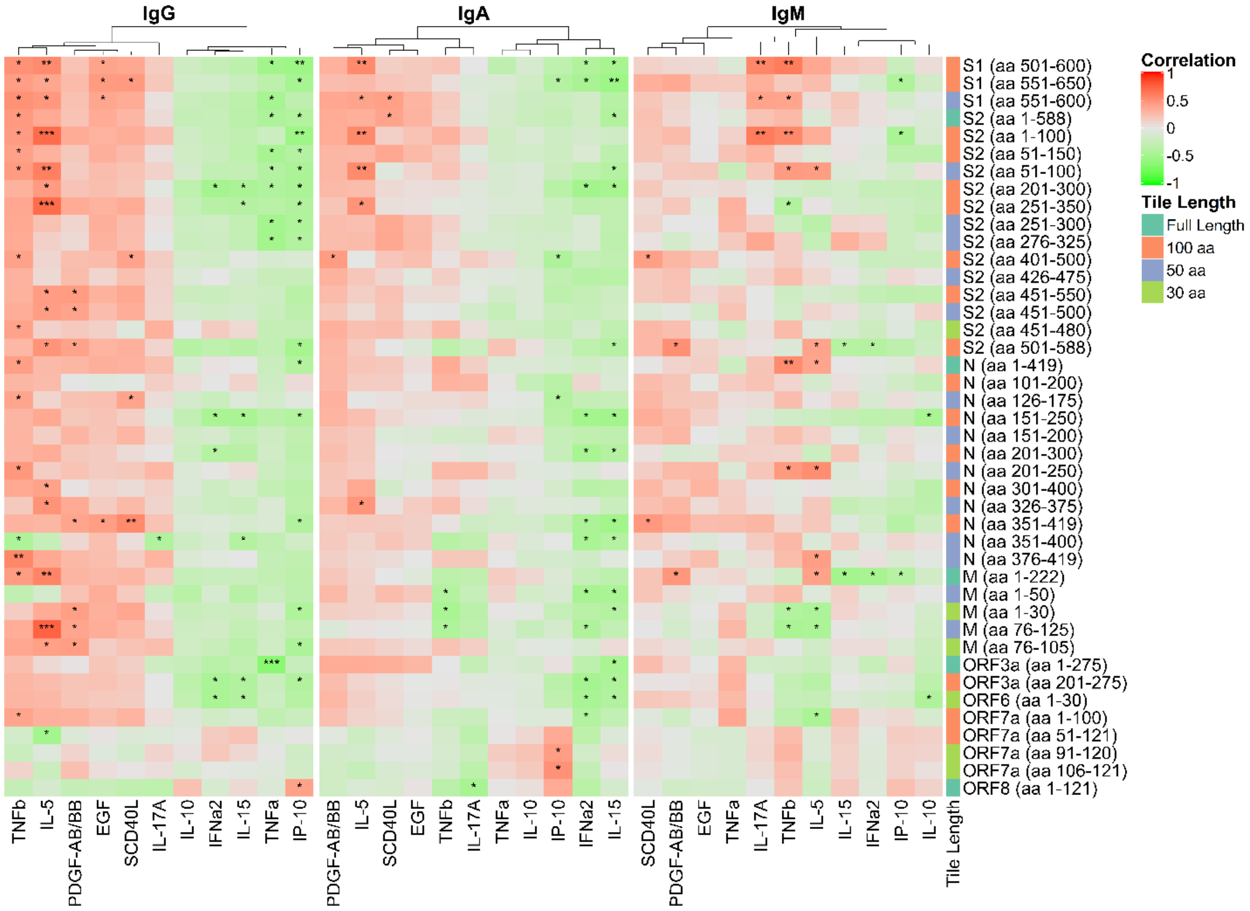


**Supplemental Figure S2. Correlation between reactive SARS-CoV-2 proteins and fragments with selected cytokine levels.** The heatmap shows the Pearson's correlation coefficient between antibody and cytokine levels on a colorimetric scale from negative correlation in green to positive correlation in red. The antigens displayed correspond to proteins and fragments produced *in vitro* that were seropositive ($\geq$ 1.0 $\log_2$ normalized signal intensity) in at least 10% of the study population (i.e. at least 3 individuals). The cytokines displayed were selected based on significant associations with antibody levels in linear mixed effects regression models, adjusted for age category, sex and requirement of ventilator. Significance of the correlations are shown by overlaid asterisks (* = p<0.05, ** = p<0.005, *** = p<0.0005). IL-17A and IL-5 were selected due to significant associations with individual antibody responses. Protein/fragment information is annotated to the right of the heatmaps and includes the protein name and the amino acid coordinates in parentheses and the length of the protein fragments ("Tile Length", as full length, 100, 50 or 30 aa).

#### Supplemental Figure S3

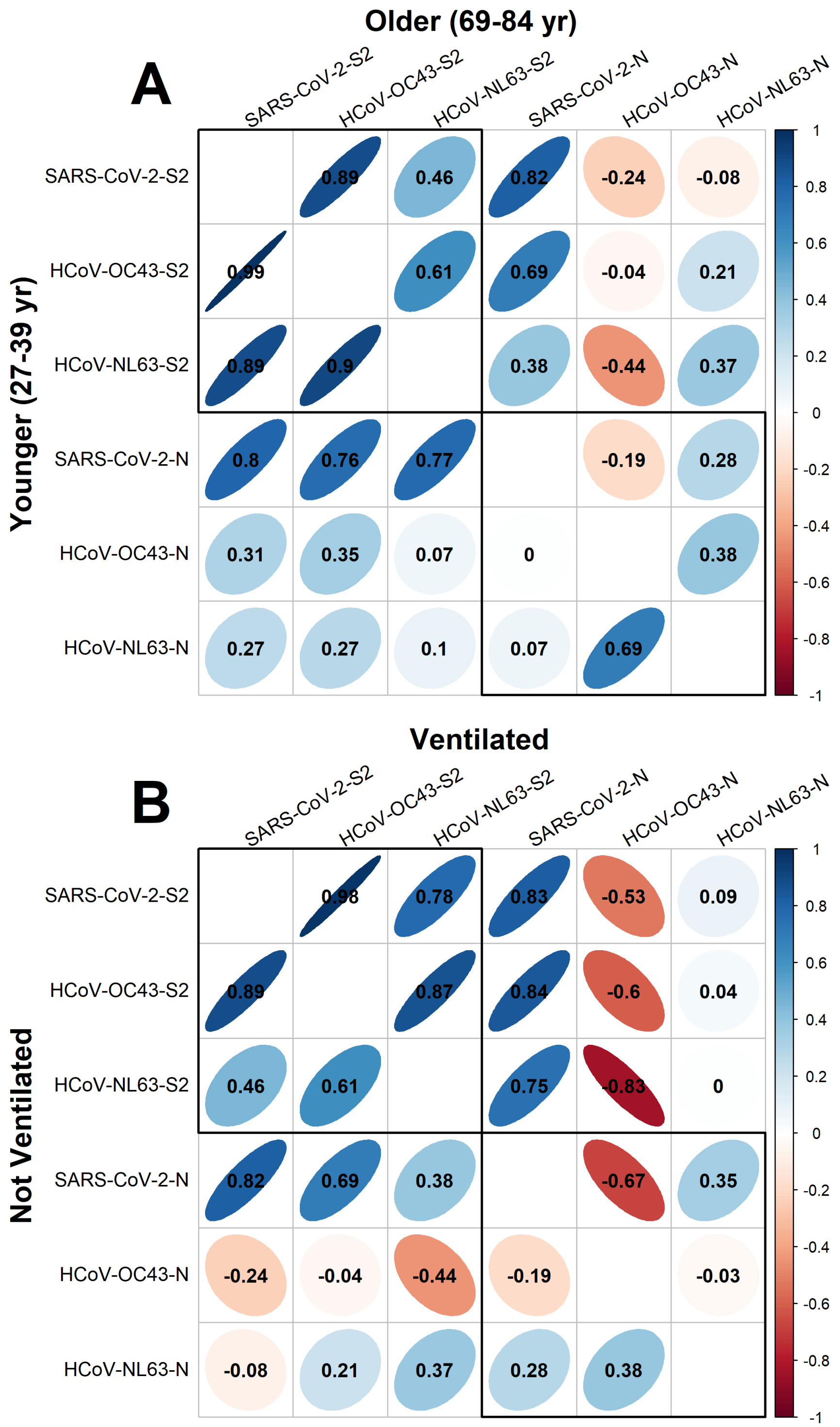


**Supplemental Figure S3. Correlation between IgG responses to SARS-CoV-2 full N and S2 proteins and other endemic human coronaviruses stratified by age and severity. (A-B)** Correlogram depicting the Pearson's correlation coefficient ($\rho$) between IgG normalized signal intensity to SARS-CoV-2, HCoV-OC43 and HCoV-NL63 N and S2 full-length proteins produced *in vitro*. In panel **A**, the lower left half of the diagonal shows correlation between reactivity of sera in the younger age group (27-39 yr), and the upper right half of the diagonal shows the older group (69-84yr). In panel **B**, the lower left half of the diagonal shows correlation between reactivity of sera in the non-ventilated group and the upper half of the diagonal shows the ventilated group. The color scale indicates positive correlation in darker shades of blue and negative correlation in darker shades of red, and $\rho$ is overlaid on each comparison. Additionally, the narrowness and slope of the ellipses represent increasing positive or negative correlation.

**Supplemental Figure S4**

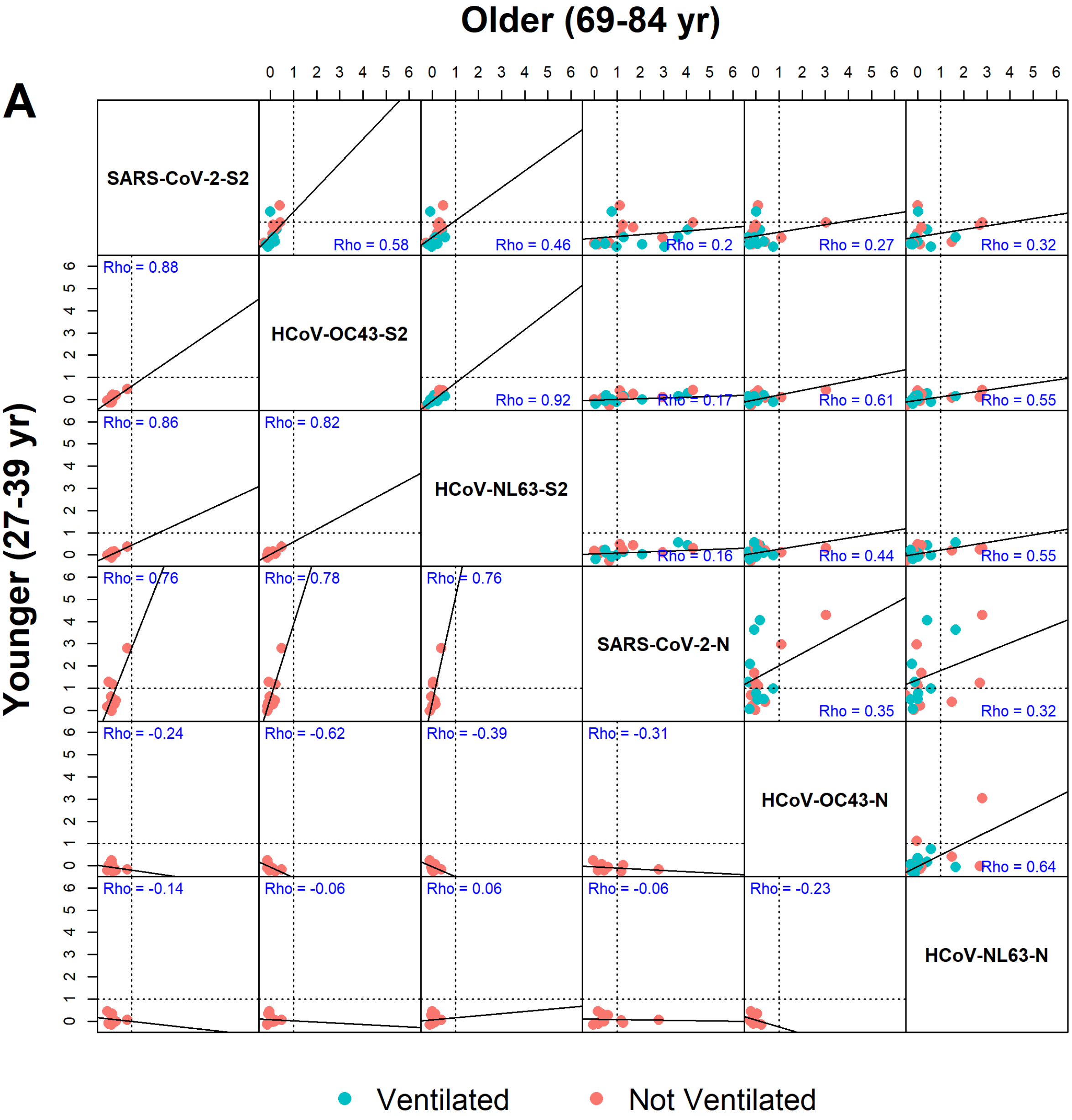

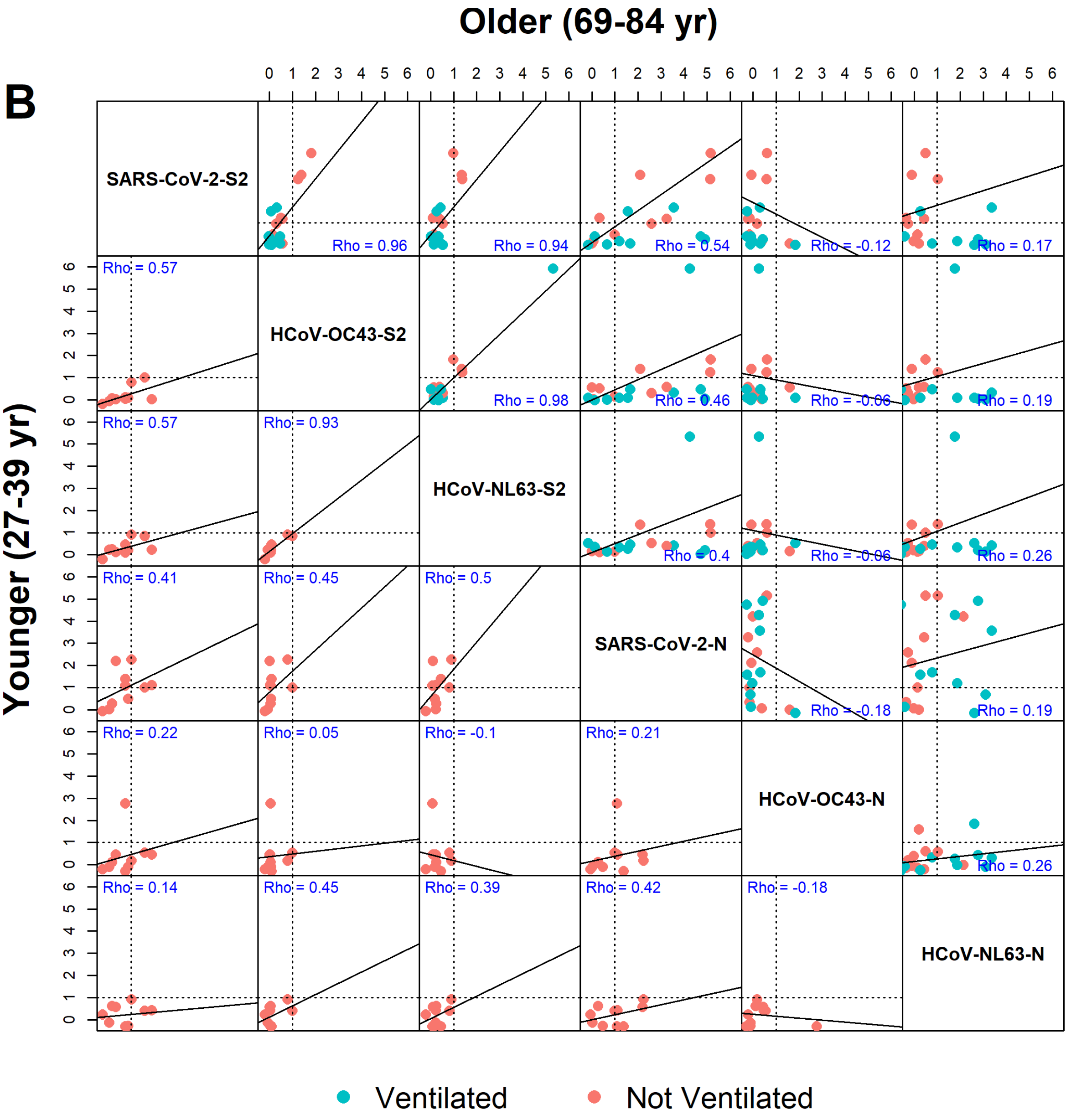


**Supplemental Figure S4. Correlation between IgA and IgM responses to SARS-CoV-2 full N and S2 proteins and other endemic human coronaviruses. (A-B)** Correlogram depicting the Pearson's correlation coefficient (*Rho*) between IgM **(A)** and IgA **(B)** normalized signal intensity to SARS-CoV-2, HCoV-OC43 and HCoV-NL63 N and S2 full-length proteins produced *in vitro*. The lower left half of the diagonal shows correlation between reactivity of sera in the younger age group (27-39 yr), and the upper right half of the diagonal shows the older group (69-84 yr). Ventilated patients are represented by teal dots and non-ventilated patients are represented by red dots. Line of seropositivity defined as a $\log_2$ normalized signal intensity ≥ 1 is depicted by horizonal and vertical dotted lines in each scatterplot. Rho coefficient listed in blue lettering in each box.

**Supplementary Figure S5**

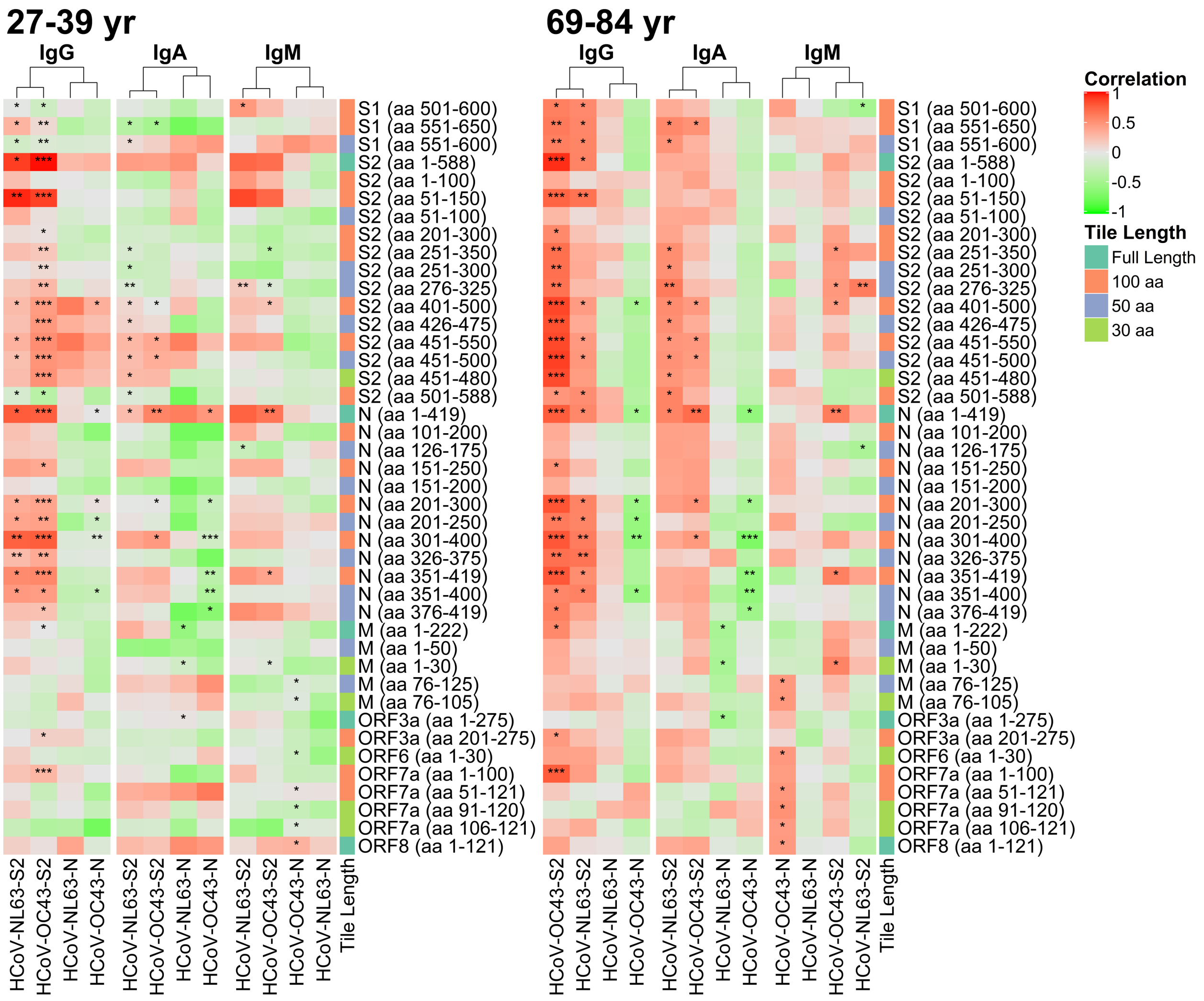


**Supplementary Figure S5. Correlation of antibody response to SARS-CoV-2 fragments and endemic coronaviruses stratified by age.** Heatmap depicting Pearson's correlation coefficients between antigenic antibody fragments and endemic coronaviruses on a colorimetric scale from negative correlation in green to positive correlation in red. The antigens displayed correspond to proteins and fragments produced *in vitro* that were seropositive (≥ 1.0 $\log_2$ normalized signal intensity) in at least 10% of the study population (i.e. at least 3 individuals). Significance of the correlations are shown by overlaid asterisks (* = p<0.05, ** = p<0.005, *** = p<0.0005). Heat maps are stratified by younger (27-39yr, left) and older age group (69-84yr, right), and by IgG, IgA and IgM reactivities (sub panel from left to right within each age group). SARS-CoV-2 fragments are compared to full length S2 and N from HCoV-OC43 and HCoV-NL63. Protein/fragment information is annotated to the right of the heatmaps and includes the protein name and the amino acid coordinates in parentheses and the length of the protein fragments ("Tile Length", as full length, 100, 50 or 30 aa).

#### Supplementary Figure S6

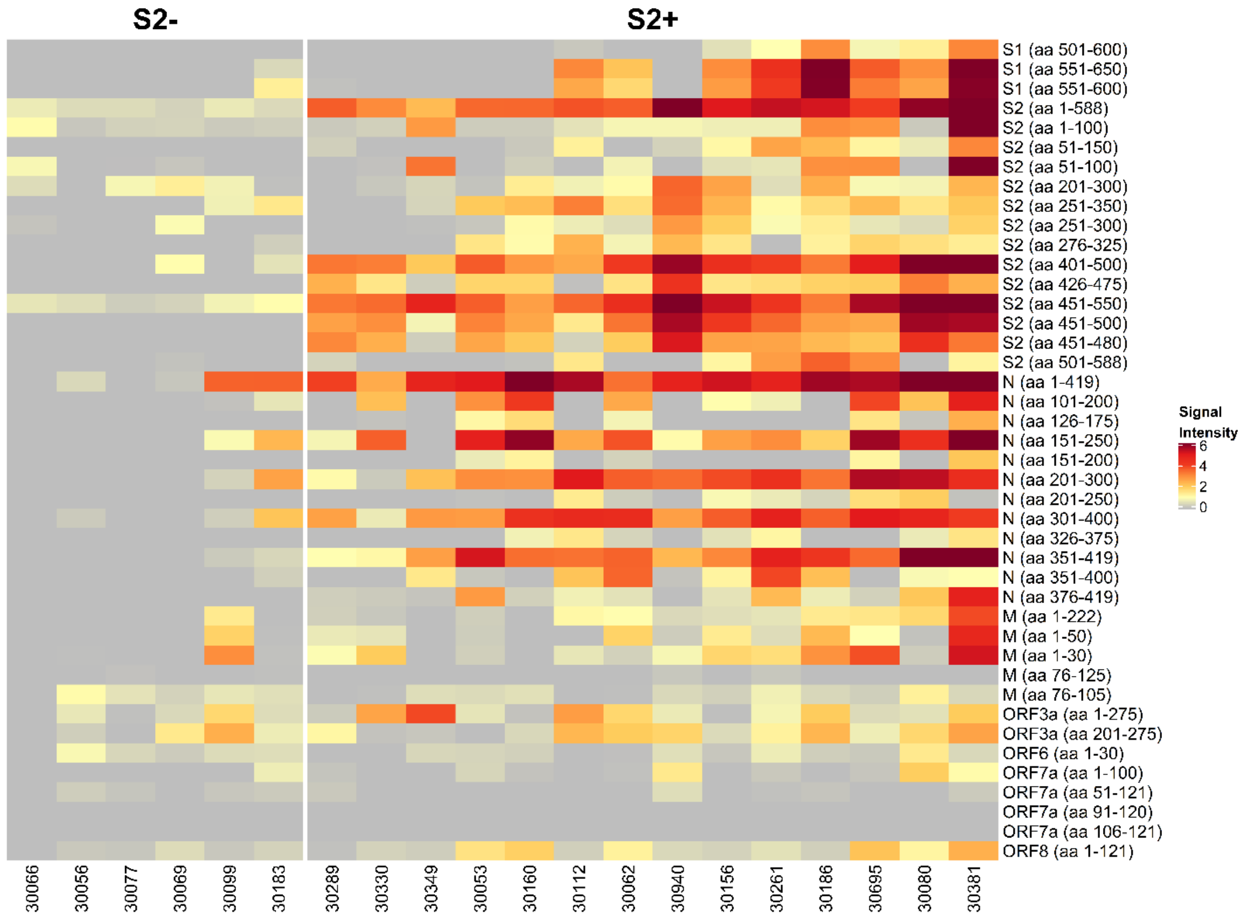


**Supplementary Figure S6. Antibody response to SARS-CoV-2 fragments in older adult patients separated by seroreactive and serosilent patients to full length S2 fragment.** Heatmap presenting the signals of antibody binding to individual protein fragments produced *invitro* within the antigenic regions of SARS-CoV-2 separated by patients that are seroreactive to full S2 protein produced *invitro* (S2+) and those that are seronegative to full length S2 protein produced *invitro* (S2-). Columns represent serum samples within the older adult age group. Antibody $\log_2$ signal intensity is shown on a color scale from grey to red. Fragments displayed correspond to proteins and fragments produced *in vitro* that were seropositive ($\geq$ 1.0 $\log_2$ normalized signal intensity) in at least 10% of the study population.
